# Testing the internal validity of the maintenance/discontinuation study design for antidepressant medications in depression using simulations

**DOI:** 10.1101/2025.10.02.25337182

**Authors:** William U. Meyerson, Daniel Schäfer, Ashley Dankese, Tianxi Cai, Jordan W. Smoller

## Abstract

**Background:** Evidence for longer-term antidepressant use in major depressive disorder comes largely from maintenance/discontinuation trials. Among patients who remit on antidepressants, approximately 20% relapse within one year when treatment is maintained, compared with 40% when treatment is discontinued. This apparent benefit exceeds what might be expected from the modest effects observed in acute-phase trials, raising questions about the internal validity of maintenance/discontinuation trials. By randomizing only patients who improve during initial treatment, these trials may enrich for patients who benefit from medication. Whether this enrichment is sufficient to explain the observed maintenance benefit remains unclear.

**Objective:** To test whether enrichment can account for the apparently larger treatment effects in maintenance/discontinuation trials.

**Methods:** We simulated maintenance/discontinuation trials using design parameters averaged across nine trials, symptom fluctuations derived from STAR*D, and drug-effect estimates from placebo-controlled acute-phase trials. We compared simulated relapse rates and relative risks with aggregated post-randomization trial outcomes that were withheld from model calibration.

**Results:** Selection of remitters increased the simulated average drug benefit 1.7-fold, from 1.75 Hamilton Depression Rating Scale points before selection to 2.91 points among participants entering the randomized phase. Simulated relapse rates in both arms and relative risks showed excellent agreement with observed trial outcomes according to the study’s model-fit criterion.

**Conclusions:** Under the modeled assumptions, enrichment can account for the larger treatment effects observed in maintenance/discontinuation trials. These findings support the internal validity of these trials for estimating the benefits of continued antidepressant treatment among patients who remitted on medication.

## Introduction

An estimated 8.8 million Americans have been taking antidepressant medications (ADMs) continuously for 10 or more years^1^. Clinical guidelines recommend that ADMs be continued for at least 6-12 months after remission of an episode of major depressive (MDD) to prevent relapse, and for longer periods in individuals with 3 or more lifetime episodes^2,3^.

The evidence base for this practice does not come from classical acute phase randomized controlled trials (RCTs), because in MDD these are rarely longer than 3 months^1,4^. Instead, the recommendations derive from maintenance/discontinuation trials (M/D trials), in which participants are treated with ADM in an open-label fashion in phase 1, after which responders are randomized in phase 2 to continued ADM vs placebo substitution. Meta-analyses of M/D trials report that relapse rates are 37%-45% after 6-12 months in discontinuation arms but only 18%-24% in the continuation arms ^5–9^.

When appropriate for the condition and treatment and appropriately conducted, M/D trials have been praised as an ethical and efficient alternative to classical acute-phase randomized controlled trials, especially when longer follow-up times are desired^10^. However, there is a lack of consensus about whether the M/D design is internally valid in the specific case of ADMs for MDD^11–13^. Critics have noted the puzzling discrepancy between the typically modest effect sizes of ADMs in acute phase RCTs (approximately 1.8 points on the 17-item Hamilton Depression Rating Scale per one authoritative meta-analysis)^14^ and the frequently larger benefit of ADM treatment in M/D trials.

A leading explanation for this discrepancy points to selection effects. Because only phase 1 responders enter phase 2, the latter is enriched for patients who can tolerate ADMs and who receive benefit from them. Because of this enrichment, it would be inappropriate to directly extrapolate the results of M/D trials to patients whose acute response was not sufficient to make them eligible for the randomized phase. Nevertheless, provided the trials are internally valid for the selected cohort, their conclusions remain applicable to clinically similar patients, who comprise a substantial share of real-world practice.

While there are good reasons to believe that selection effects operate in M/D trials, it is less clear whether these effects are strong enough to fully explain the large difference in outcomes between arms in M/D trials. For example, because the natural history of depression is highly variable and responsive to placebo effects, it is unclear just how successfully the M/D design enriches for patients who benefit specifically from drug. There are other possible explanations for the effect sizes of M/D trials (e.g. withdrawal-relapse confusion^15^, discontinuation-enhanced unblinding^16,17^, rebound/”payback”^18^) which challenge the internal validity of M/D trials. So long as it remains unknown whether selection effects are sufficient to explain the results of M/D trials, the validity of practice guidelines derived from them may be questioned.

To address this gap, we performed simulations to examine whether selection effects are sufficient to explain the between-arm differences in relapse rates seen in M/D trials. The simulations are run with parameters sourced from the pre-randomization phase of recent M/D trials, together with information about average treatment effect and estimated distribution of treatment effect from acute antidepressant RCTs. These simulations test whether the results of M/D trials are consistent with what is known about the magnitude of ADM benefits from acute-phase RCTs; symptom fluctuations in treated MDD from longer-term, non-placebo-controlled studies; and the unique aspects of the study design of M/D trials. As noted, these simulations have implications for the internal validity of M/D trials of ADMs in MDD and for clinical guidelines about maintenance treatment.

## Methods

### Overview of model

The simulation model consists of 2 phases and 4 effects. Phase 1 is the single-armed open-label treatment phase. Phase 2 is a two-arm maintenance/discontinuation phase. The four effects driving the potential outcome under the simulation models are 1) symptom fluctuations derived from treated depression as a base layer in all phases and arms, 2) trial-specific placebo effects due to “contextual factors”^19^as an additional layer in all phases and arms, 3) removal of active drug effects in the discontinuation arm of Phase 2, and 4) the loss of open-label certainty in both arms once patients enter the randomized phase.

In each run of the simulation, a simulated initial cohort meeting eligibility criteria experiences a change in depression severity during open-label treatment (phase 1 related to symptom fluctuations derived from treated depression and empirical trial-specific random-effects. Participants who remit then enter phase 2, where some proportion are randomly assigned to continue antidepressant and the rest are switched to placebo. At that point, participants in the discontinuation arm lose whatever benefit they experienced from active drug according to a decay curve, and participants in both arms lose a portion of placebo expectancy effects. Relapse survival curves by arm in phase 2 are calculated according to a relapse rule (Table 1) and compared with relapse rates from real M/D trials. Results were generated across 100 runs.

**Table 1:** Characteristics of included studies.

| Study | Agent | P1 N | P2 N | P1 weeks | P2 weeks | Min DSEV (HAMD) | Remit Rule* (HAMD) | Relapse Rule* (HAMD) |
|---|---|---|---|---|---|---|---|---|
| Thase_2022<sup>20</sup> | Vortioxetine | 1105 | 580 | 16 | 32 | 20 | <=10 | >=17 |
| Durgam_2019<sup>21</sup> | Levomilnacipran | 644 | 324 | 20 | 26 | 20 | <=10 | >=15 |
| Durgam_2018<sup>22</sup> | Vilazodone | 1204 | 564 | 20 | 28 | 20 | <=10 | >=15 |
| Shiovitz_2014<sup>23</sup> | Levomilnacipran | 734 | 345 | 12 | 24 | 17 | <=10 | >=17 |
| Rosenthal_2013<sup>24</sup> | Desvenlafaxine | 874 | 548 | 20 | 26 | 20 | <=11 | >=16 |
| Goodwin_2013a<sup>25</sup> | Agomelatine | 551 | 367 | 8 | 24 | 22 | <=10 | >=16 |
| Goodwin_2013b<sup>25,28</sup> | Agomelatine | 492 | 339 | 10 | 24 | 22 | <=10 | >=16 |
| Boulenger_2012<sup>26</sup> | Vortioxetine | 639 | 396 | 12 | 24 | 20 | <=11 | >=16 |
| Rickels_2010<sup>27</sup> | Desvenlafaxine | 594 | 375 | 12 | 24 | 20 | <=8 | >=17 |

P1=Phase 1 (open-label), P2=Phase 2 (maintenance/discontinuation phase), DSEV=depression severity, HAMD=depression severity in units of the 17-item Hamilton Depression Rating Scale, after converting alternative scales to published linkage formulae (Stone 2023). \*Multiple studies had additional criteria for remission or relapse such as Clinical Global Impression threshold or the use of repeat testing. The detailed rules for converting these to a single threshold are described in the Supplementary Appendix.

### Identification of Maintenance/Discontinuation Trials to Simulate

M/D studies were included in our trial pool if they met the following criteria: 1) they were placebo-controlled M/D studies of antidepressants for MDD included in the most recent systematic review on the topic^6^; 2) no treatment arm included psychotherapy as a study intervention; and 3) a report of the study was published within 15 years of manuscript preparation (i.e. since 2010). Nine RCTs^20–27^ met these inclusion criteria with selected characteristics listed in Table 1.

### MODEL Parameters

Model parameters are listed in Table 2. The number of participants in phase 1, target number of participants in phase 2, duration of phase 1 and 2 in months, the minimum depression score for eligibility, remission threshold, relapse threshold, minimum duration of current episode, maximum duration of current episode, and proportion of participants assigned to continue drug during phase 2 were calculated as the mean value of these variables from the nine included trials, and rounded to the nearest integer where appropriate. The lifetime number of depressive episodes, and maximum age for study inclusion were obtained from the median of these values among the included studies. Most included studies used a compound remission rule, where participants must also meet some less strict cutoff at an intermediate time-point during phase1; on average this cutoff was less strict than the end-phase-1 remission threshold by 2 points. To account for this, we implemented a less-strict cutoff at month 2 in the simulations, which participants had to pass before reaching the more-strict cutoff at month 3.

**Table 2:** Trial-Averaged Parameter Set Used for Simulation.

| Parameter | Base Case Value | Sensitivity Analysis Values | Reference |
|---|---|---|---|
| Number of participants in Phase 1 | 760 | 492, 1204 | § |
| Target fraction of participants in Phase 2 | 0.59 | 0.47, 0.69 | § |
| Phase 1 duration (months) | 3 | 2,4 | § |
| Phase 2 duration (months) | 6 | 5,7 | § |
| Eligibility*: Minimum Depression score (HAM-D) | 20 | 17,22 | § |
| Eligibility: Minimum Number of prior depressive episodes | 3 | 2, 4 | § |
| Remission threshold in last phase 1 month<br>(HAM-D) | 10 | 8, 11 | § |
| Relaxation of remission threshold in penultimate phase 1 month<br>(HAM-D) | +2 | +0, +4 | § |
| Proportion randomized to continue drug | 0.57 | 0.49, 0.74 | § |
| Proportion of participants who benefit from drug | 0.23 | 0.115, 0.46 | <sup>14</sup> |
| True drug effect in those who benefit from drug<br>(HAM-D) | -1.75 | -1.86, -1.63 | <sup>14</sup> |
| Percentage of drug effect lost 1 month after discontinuation | 65% | 50%, 75% | <sup>29</sup> |
| Percentage of drug effect lost 2 months after discontinuation | 85% | 75%, 100% | <sup>29</sup> |
| Expectancy placebo effect when 100% are administered drug | -1.43 |  | <sup>30</sup> |
| Study-specific common factors<br>(HAM-D) | -1.9 | Calculated each run | § |
| Remission threshold (HAM-D) | 16 | 15, 17 | § |

\*Additional eligibility criteria: No co-morbid Axis I or II disorder, age between 18 and 70, duration of current depressive episode between 35 and 570 days. § = The nine included RCTs.

The average drug treatment effect was obtained from a recent meta-analysis of acute-phase RCTs^14^. The supplement describes how we used other data from this source to characterize the distribution of this effect among participants. In the absence of additional information, we assumed that the benefits of active drug wash out after discontinuation according to a decay curve that is the mirror image of the onset curve of antidepressant efficacy, obtained from a meta-analysis of the time-course of onset of ADM effects^29^.

The magnitude of the expectancy effect as a function of the proportion of participants assigned to drug was obtained from a meta-analysis of antidepressant RCTs with differing randomization ratios^30^.

The contextual factors effect was a constant value added to the depression severity scores of all participants at all follow-up time-points, that was chosen through a parameter grid search to result in the number of phase 1 participants meeting remission to be as similar as possible to the number in the included trials.

### Natural History of Treated Depression

Sequenced Treatment Alternatives to Relieve Depression (STAR-D) was the largest long-term publicly funded trial of antidepressants in MDD and is the field standard for characterizing the natural history of treated depression^31^. Our simulations made use of the depression trajectories of STAR-D participants in months 0 through 12 as a base layer to capture realistic month-to-month fluctuations of depression severity and measurement noise, with other features of the simulation layered on top of this base layer. Drug effects were modeled separately using estimates from placebo-controlled acute-phase trials. We assumed that the symptom fluctuations observed in treated STAR-D participants also represent those that would occur in the absence of active medication.

To ensure simulations represented similar patients as the RCTs, we excluded STAR-D participants with a comorbid psychiatric disorder to better reflect the M/D RCT data. Likewise we only included STAR-D participants who met the composite M/D trial thresholds for age, baseline depression severity, depressive episode history, and duration of the current episode (Table 2).

To ensure sufficient sample size for matching and to complete missing data, we augmented the STAR-D data with 100 synthetically generated complete cohort replicates designed to preserve relationships among variables. This was achieved through a Bayesian hierarchical local mixture model with local dependence^32^ implemented via the *multipleSyn* function from synMicrodata^33^ R package. (v2.1.0), with a burn-in of 100, an interval of 50 between synthesis steps, and otherwise default parameters. As a post-processing step, depression severities in months 7-12 -months with high (69.0%-80.4%) missingness in the original data-were normalized to have a stable mean depression severity concordant with lower-missingness (62.1%) month 6.

The synthetic data had high fidelity to the original data. For example, the difference in depression severity between months 0 and 1 had a mean (SD) of 8.3 (7.5) in the original data and 8.3 (9.0) in the synthetic data, and the correlation between the number of prior episodes and depression severity in month 6 was 0.085 in the original data and 0.064 in the synthetic data.

### Model Scoring

We evaluated model fit by comparing simulated outcomes to withheld post-randomization results averaged across the nine included trials. At the end of each post-randomization month (months 1–6), we assessed three outcomes: (1) cumulative relapse frequency in the continuation arm; (2) cumulative relapse frequency in the discontinuation arm; and (3) the relapse relative risk (continuation vs. discontinuation).

Among the nine included trials, the mean and standard deviation of these 3 outcome types were calculated at each of these 6 time points, after extracting study-specific relapse rates via plot digitization (graphreader.com) (Supplemental Table S3). One study (Shiovitz 2014) lacked data for the 6-month mark, which was imputed using the *mice* R package (v3.17.0).

The goal of this project was to develop a general sense of whether the results of M/D trials are broadly consistent with what we know about MDD and antidepressants from short-term acute phase RCTs, non-placebo-controlled longer studies, and the study design of M/D trials. To support this goal, we used the following pragmatic scoring system. For each outcome and time point, we computed a Z-score as the difference between the simulation mean and the mean across the nine included studies, divided by the standard deviation of that study mean. The mean absolute value of Z-scores across all outcomes and time-points was taken as a summary score. This summary score was interpreted according to rubric in Table 3.

**Table 3.** Interpretation rubric for summary score fit.

| **Summary Score** | **Interpretation** | **Comment** |
|---|---|---|
| 0-0.66 | Excellent | Here, the average departure of the model from the trial average is well below the standard deviation between individual trials. |
| 0.67-1.32 | Good |  |
| 1.33-1.99 | Fair |  |
| 2.0+ | Poor | Here, the average departure of the model from the trial average is 2 or more standard deviations away from the individual trials. |

## Results

### Main Model

#### Efficiency of enrichment

The mean HAM-D difference between the maintenance and discontinuation arms during post-randomization months 3–6 was 2.91 points, approximately 1.7 times the average treatment effect from acute RCTs used to parameterize Phase 1. This amplification is consistent with enrichment arising from restricting Phase 2 to Phase-1 responders.

#### Relative risk of relapse

The relative risk of relapse in the continuation vs discontinuation arm was 0.52 at 3 months and 0.56 at 6 months in the simulations (Figure 2; Table 4). This was similar to the corresponding values from the withheld post-randomization phase of the six positive trials (Boulenger 2012), (Durgam 2019), (Goodwin 2013 Study 2), (Rickels 2010), (Rosenthal 2013), and (Thase 2022) but a greater reduction than the negative and/or weak-effects trials of (Durgam 2018), (Goodwin 2013 Study 1), and (Shiovitz 2014). Altogether, the relative risk of relapse in the continuation vs discontinuation arm in the simulations represented a greater risk reduction than the nine-trial-average, but because of the spread between the positive trials and negative trials, this discrepancy was less than one standard deviation in magnitude. The relative risks in our simulations were even closer to the relative risks reported in meta-analyses of a larger number of M/D studies; e.g. (Kishi 2023) reports a relative risk of relapse of 0.49 (0.43 to 0.57) at 3 months and 0.51 (0.46 to 0.58) at 6 months^34^.

**Table 4:** Comparison of relapse rates by arm in simulated and observed trials.

|  | **Relapse rate in drug arm** |  |  | **Relapse rate in placebo arm** |  |  | **Relative risk in drug arm with respect to placebo** |  |  |
|---|---|---|---|---|---|---|---|---|---|
| Month | Sim | Obs | Z | Sim | Obs | Z | Sim | Obs | Z |
| 1 | 0.033 | 0.056 | -1.23 | 0.072 | 0.086 | -0.24 | 0.46 | 0.85 | -0.72 |
| 2 | 0.092 | 0.10 | -0.28 | 0.17 | 0.16 | 0.10 | 0.53 | 0.70 | -0.84 |
| 3 | 0.12 | 0.13 | -0.20 | 0.23 | 0.22 | 0.08 | 0.52 | 0.64 | -0.58 |
| 4 | 0.16 | 0.15 | 0.13 | 0.29 | 0.26 | 0.27 | 0.55 | 0.65 | -0.48 |
| 5 | 0.19 | 0.17 | 0.28 | 0.36 | 0.29 | 0.54 | 0.51 | 0.63 | -0.67 |
| 6 | 0.25 | 0.20 | 0.78 | 0.44 | 0.32 | 0.90 | 0.56 | 0.67 | -0.49 |

Sim=Relapse rate or relative risk within the simulated data set. Obs=Mean relapse rate or relative risk average across 9 real RCTs. Z=the Z-score of the difference between the simulated value and the spread of the corresponding 9 real RCT values.

#### Continuation arm relapse rates

The absolute rates of relapse in the continuation arm were generally concordant with the relapse rates among the trials. This was particularly true at the 3-month mark, where the 11.9% cumulative relapse rate in the simulations was very similar to the nine-trial-average of 12.7%. The simulated continuation arm relapse rates somewhat undershot the nine-trial-average in months 1 and 2, and overshot them in month 4-6, but the only such month that departed from the nine-trial-average by more than 1 SD was month 1, with a relapse rate in the simulations of 3.3% vs a nine-trial-average of 5.2%; in comparison, (Thase 2022) had a 3.4% relapse rate in the drug arm at month 1.

#### Discontinuation arm relapse rates

The absolute rate of relapse in the discontinuation arm were also generally concordant with the relapse rates among the trials. In the simulations, cumulative relapse rates ranged from 7.2% at month 1 to 43.9% at month 6; which paralleled a nine-trial-average that ranged from 8.6% at month 1 to 31.5% at month 6, although tending to overshoot the nine-trial-average in the later months.

#### Model scoring

Across the 3 outcomes and 6 time points, the mean departure of the simulation from the nine-trial-average was 0.49 standard deviations, representing excellent fit according to the interpretation schema of Table 3. Of the 18 outcome-time pairs, 12 met criteria for excellent fit, 6 met criteria for good fit, and none met criteria for fair or poor fit.

**Figure 1.**
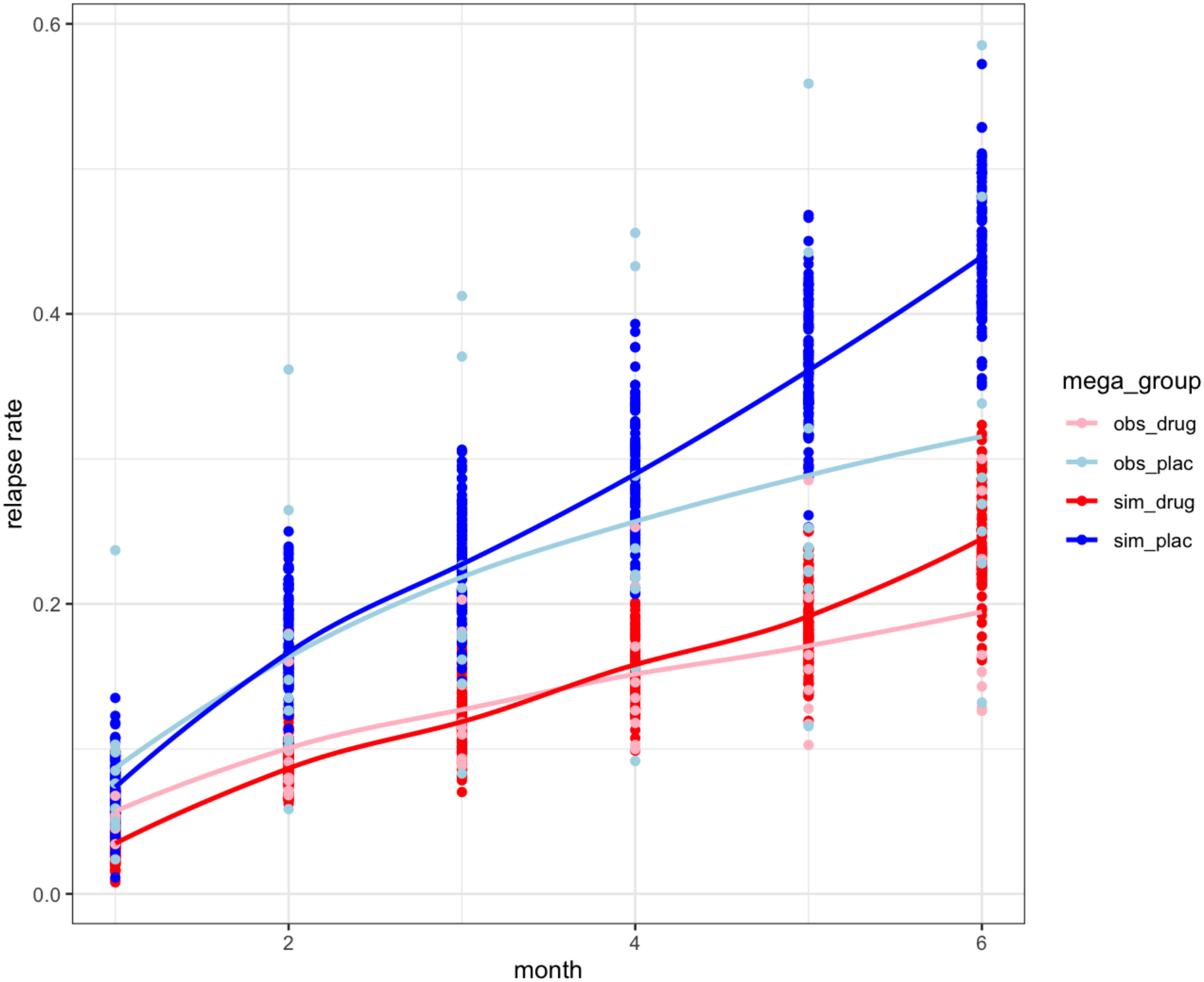
Simulated versus observed relapse rates by arm in simulations and real RCTs.

## Discussion

Maintenance/discontinuation trials form the bedrock of evidence for the common practice of continuing antidepressant medications, but it is unclear whether their large effect sizes are consistent with the modest effects of ADMs in acute-phase RCTs. Our simulations demonstrate that typical relapse curves of continuation and discontinuation arms on ADM M/D trials are a straightforward extension of what we already know from acute phase studies, the natural history of depression, and the design of M/D trials. Our simulations had excellent fit to real trial data for all three outcome types: maintenance arm relapse rates, discontinuation relapse rates, and relative risk of relapse between arms. The large risk reduction in the maintenance arm is expected to be a consequence of the 1.7-fold enrichment of the average treatment effect that occurred after selection of remitters during phase 1 of the simulation. These results do not imply that withdrawal-relapse confusion, unblinding, and rebound do not occur in M/D trials, only that these phenomena are not necessary to explain the basic results of the ADM M/D trials.

Interestingly, the relative risk of relapse in the simulations hewed even more closely to meta-analyses of a larger number of studies than to the sampling of nine recent studies on which the simulations were based^34^. Some possible explanations for this finding include heterogeneity in patient population at the time of treatment initiation or difference in the details of study procedure or execution contributing to high “random effects” idiosyncratic variance between individual studies that was not captured in our simulations, but that they may be triangulating a more general truth consistent with what is known from ADM acute phase RCTs.

One limitation of this work is the inclusion of only M/D trials from the last 15 years. Moreover, the STAR-D data used to model natural history carries a number of limitations including the fact that participants’ treatment was switched, although we believe the impact of this is contained by the fact that no treatments in STAR-D were found to be superior to any other^35^. The simulations were not precise enough to incorporate nuances of remission and relapse rules such as stability between consecutive weeks. Some model parameters carry high uncertainty such as in how antidepressant effects are distributed, with different authors proposing different degrees of heterogeneity^14,36^

Nonetheless, to the extent that these simulations capture the essential features of ADMs and MDD, these results increase our confidence that M/D trials are internally valid and therefore strengthen the evidence supporting clinical guidelines that draw on these trials when discussing maintenance treatment for patients who remitted on antidepressant medication.

## Data Availability

No new data were generated for this study.
The STAR*D data can be obtained from the NIMH Data Archive, with more information in the Acknowledgments section.
The RCT summary data are publicly available from the cited trials.

## Acknowledgments

Data and/or research tools used in the preparation of this manuscript were obtained from the National Institute of Mental Health (NIMH) Data Archive (NDA). NDA is a collaborative informatics system created by the National Insritutes of Health to provide a national resource to support and accelerate research in mental health. Dataset identifier(s): 10.15154/xx52-jp36 and 10.15154/zk9m-5n40. This manuscript reflects the views of the authors and may not reflect the opinions or views of the NIH or of the Submitters submitting original data to NDA.

## Author Contributions

Dr Meyerson had full access to all of the data in the study and takes responsibility for the integrity of the data and the accuracy of the data analysis.

*Concept and design:* Meyerson

*Acquisition, analysis, or interpretation of data:* Meyerson, Schäfer, Dankese

*Drafting of the manuscript:* Meyerson

*Critical review of the manuscript for important intellectual content:* All authors.

*Statistical analysis:* Meyerson

*Obtained funding:* Meyerson, Smoller

*Administrative, technical, or material support:* Smoller

*Supervision:* Cai, Smoller

